# NeMoR: a New Method Based on Data-Driven for Neonatal Mortality Rate Forecasting

**DOI:** 10.1101/2021.04.22.21255916

**Authors:** Carlos Eduardo Beluzo, Luciana Correia Alves, Natália Martins Arruda, Cátia Sepetauskas, Everton Silva, Tiago Carvalho

## Abstract

Reduction in child mortality is one of the United Nations Sustainable Development Goals for 2030. In Brazil, despite recent reduction in child mortality in the last decades, the neonatal mortality is a persistent problem and it is associated with the quality of prenatal, childbirth care and social-environmental factors. In a proper health system, the effect of some of these factors could be minimized by the appropriate number of newborn intensive care units, number of health care units, number of neonatal incubators and even by the correct level of instruction of mothers, which can lead to a proper care along the prenatal period. With the intent of providing knowledge resources for planning public health policies focused on neonatal mortality reduction, we propose a new data-driven machine leaning method for **N**eonatal **M**ortality **R**ate forecasting called **NeMoR**, which predicts neonatal mortality rates for 4 months ahead, using **NeoDeathForecast**, a monthly base time series dataset composed by these factors and by neonatal mortality rates history (2006-2016), having 57,816 samples, for all 438 Brazilian administrative health regions. In order to build the model, Extra-Tree, XGBoost Regressor, Gradient Boosting Regressor and Lasso machine learning regression models were evaluated and a hyperparameters search was also performed as a fine tune step. The method has been validated using São Paulo city data, mainly because of data quality. On the better configuration the method predicted the neonatal mortality rates with a Mean Square Error lower than 0.18. Besides that, the forecast results may be useful as it provides a way for policy makers to anticipate trends on neonatal mortality rates curves, an important resource for planning public health policies.

**Graphical Abstract:** 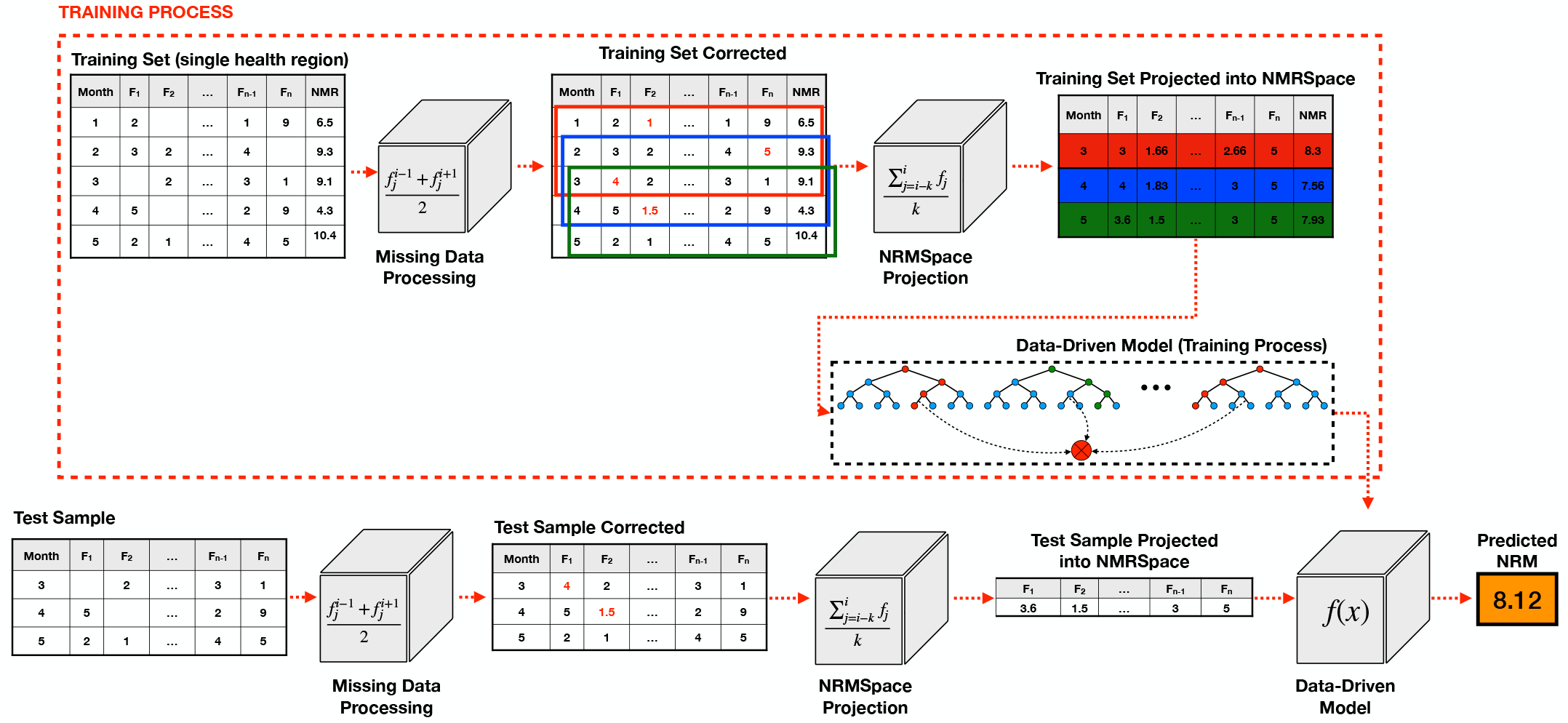

**Highlights:**

- Proposition of a new data-driven approach for neonatal mortality rate forecast, which provides a way for policy-makers to anticipate trends on neonatal mortality rates curves, making a better planning of health policies focused on NMR reduction possible;
- a method for NMR forecasting with a MSE lower than 0.18;
- an extensive evaluation of different Machine Learning (ML) regression models, as well as hyperparameters search, which accounts for the last stage in NeMoR;
- a new time series database for NMR prediction problems;
- a new features projection space for NMR forecasting problems, which considerably reduces errors in NRM prediction.

## 1. Introduction

Neonatal Mortality Rate (NMR) and Infant Mortality Rate (IMR) are strong indicators to measure the quality of public health systems, living conditions and development level of a country, which makes their proxy of the poverty and socioeconomic conditions [9]. Rates of perinatal and neonatal mortality reflect factors related to the quality of prenatal and childbirth care, while the postnatal component is mainly related to socio-environmental factors [12].

A reduction in child mortality rate is one of the United Nations (UN) Sustainable Development Goals for 2030 [19] and was also listed as one of UN’s Millennium Development Goals for 2015, which aimed to reduce Child Mortality Rates by two-thirds between 1990 and 2015.

Despite the global infant mortality rate declined from about 65 deaths per 1,000 live births in 1990 to less than 30 by 2018, this result is still far from the rate considered acceptable [20, 21].

Since the last decades of the past century, Brazil has experienced rapid demographic and epidemiological transitions. Between 1991 and 2010, the infant mortality rate dropped to 16.4 deaths per 1,000 live births and life expectancy at birth increased from about 50 to about 73 years over the same period [25]. In this transition process of mortality in Brazil, we can highlight the reduction in infant mortality for infectious and parasitic diseases, which are main risk factors associated with better life and sanitary conditions, hygiene, nutrition, access and health care Beluzo *et. al*. [1].

Despite the overall reduction in infant mortality, this occurs unevenly in different regions of the country. Moreover, one of the challenges for Brazil is the reduction of neonatal mortality, which accounts for 2/3 of infant mortality and which has been decreasing at a much slower rate than that of post-neonatal mortality [29]. The neonatal mortality rate declined from 16.7 per 1,000 in 2000 to 11.1 per 1,000 in 2010 [26].

According to the World Health Organization (WHO), preterm birth, intrapartum-related complications (birth asphyxia or lack of breathing at birth), perinatal infections, maternal factors and birth defects cause most neonatal deaths [18].

Studies conducted by Nascimento *et. al*. [16] and Migoto *et. al*. [14], have shown that some factors are strongly related to total neonatal mortality and early neonatal mortality. It was observed that neonatal deaths were related mainly to the quality of the prenatal care and direct care labor. These features were measured through some variables such as: number of prenatal consultations, type of labor, professional responsible for the childbirth (doctor on call, obstetrician, nurse or other). In addition, some associations were found regarding the reproductive history of the mother, such as if the mother presented fetal losses in previous pregnancies. They also identify some relation with the presence of malformation and to maternal socioeconomic conditions (mother’s education, marital status and mother’s race).

In a proper health system, the effect of some of these factors could be minimized by the appropriate number of newborn intensive care units, number of health care units, number of neonatal incubators and even by the correct level of instruction of mothers, which can lead to a proper care along prenatal period.

Furthermore, reduction of neonatal mortality is an essential part of the third Sustainable Development Goal (SDG), to prevent child deaths. However, in order to achieve this goal it is necessary to have a deep understanding of the levels and trends in neonatal mortality.

In this paper, based on the scientific hypotheses that combinations of different factors, including previous NMR itself, can lead to a good prediction of NMR, we proposed a new data-driven method for **N**eonatal **M**ortality **R**ate forecasting called **NeMoR**. NeMoR predicts neonatal mortality rates in a defined window of *n* months ahead using as input 5 features: (1) NMR, (2) number of health care units, (3) number of neonatal incubators in region, (4) number of mothers with high scholar level (from 8 to 11 education years) in the region, and (5) number of newborn intensive care units in the region.

In contrast to classical statistical models, NeMoR relays on knowledge learned from an extensive database, grouping more than 28,210,881 records from Brazil into health regions between 2006-2016, which generate a time series of 10 years, with monthly intervals.

**To the best of our knowledge, there are no data-driven models constructed from Brazilian data to forecast NMR by health region. As well, there are no reported results for this kind of problem using features from Mortality Information System (Sistema de Informação de Mortalidade - SIM) and the National Information System on Live Births (Sistema de Informação de Nascidos Vivos - SINASC). Then, the main contributions of this paper are: (1) proposition of a new data-driven approach for NMR forecast, which provides a way to policy-makers anticipate trends on NMR curves, making possible a better planning of health policies focused on NMR reduction; (2) an extensive evaluation of different Machine Learning (ML) regression models, as well as hyperparameters search, which accounts for the last stage in NeMoR; (3) a new time series database for NMR prediction problems; (4) a new features projection space for NMR forecasting problems, which considerably reduces errors on NRM prediction; (5) a method for NMR forecasting with a MSE lower than 0.18**.

The next sessions of this paper are organized as follows: in Section 2, we present some of the state-of-the-art methods that apply time series analysis for the prediction of different features in health problems; Section 3 presents details for methodology in dataset construction. In Section 4, we present details about NeMoR, and all its stages; Section 5 brings details of experiments performed to validate NeMoR, as well as details of results related to important choices made along method construction; finally in Section **??** we present a discussion of the experiment’s results, the main conclusions and perspectives for future works.

## 2. Time Series Forecasting Applied on Public Health Problems

When solving forecasting problems related to public health, in special the ones related to mortality rate prediction, most of the studies rely on standard methods as autoregressive integrated moving average (ARIMA) [31] and Linear Regression [4].

Shang and Smith [28], for example, applied aggregated hierarchical grouped time series by states and by gender with bootstrap method to predict counts of regional infant mortality in Australia for one-step ahead to ten-step-ahead. This method may lead to inaccurate forecasts on the top level series when data presents missing values at the bottom level of the hierarchy. They used a large period of data from 1933 to 1993 to forecast data until the end of 2003, a ten years step ahead.

Opare [17] models the under-five mortality rates (U5MR) for Ghana, comparing three different time series models: ARIMA, Bayesian Dynamic Linear Model, and Random Walk with drift models and predicting four years out of sample forecast. The authors used under-five mortality rates data from 1961 to 2012 and predicted out-of-sample from 2013 to 2016. The result presented by their study suggest that, among the three models, the Random Walk with drift model presented the best fit model for the Under-five mortality Rates for Ghana. Rotami *et. al*. [24] modeled forecast problem for monthly under-five mortality rate from 2005 to 2012 in a province in Iran using ARIMA. It predicts the impact of future variations on the U5MR and reveals that the improvement of under-five mortality data collection is necessary to achieve the best rates in Iran, so possibly the under-five mortality is under-reported. Furthermore, the authors concluded that the capability of the model to forecast in short time periods and model accuracy on predicted values were not assessed.

On Mishra *et. al*. [15], the Infant Mortality Rate (IMR) was forecasted for a period of 9 years in India, also using ARIMA. The authors used a long term period of data, 1971-2016 of infant mortality rate. Their results showed a declining trend on IMR forecasted for both sample and postsample periods. The authors concluded that despite the fact the model made a good performance on forecasting, other factors direct or indirectly impact on infant mortality. However, according to the authors, ARIMA model limitation avoids the inclusion of these factors on the model.

Forecasting models for neonatal mortality rates are restricted in developed countries. In Brazil, forecasting models to predict neonatal mortality rates are very unusual. Additionally, as can be observed on Figure 1, NMR depicts different behaviours on different health regions in Brazil^1^, which makes the generalization of NMR forecasting for the entire country very difficult.

**Figure 1:**
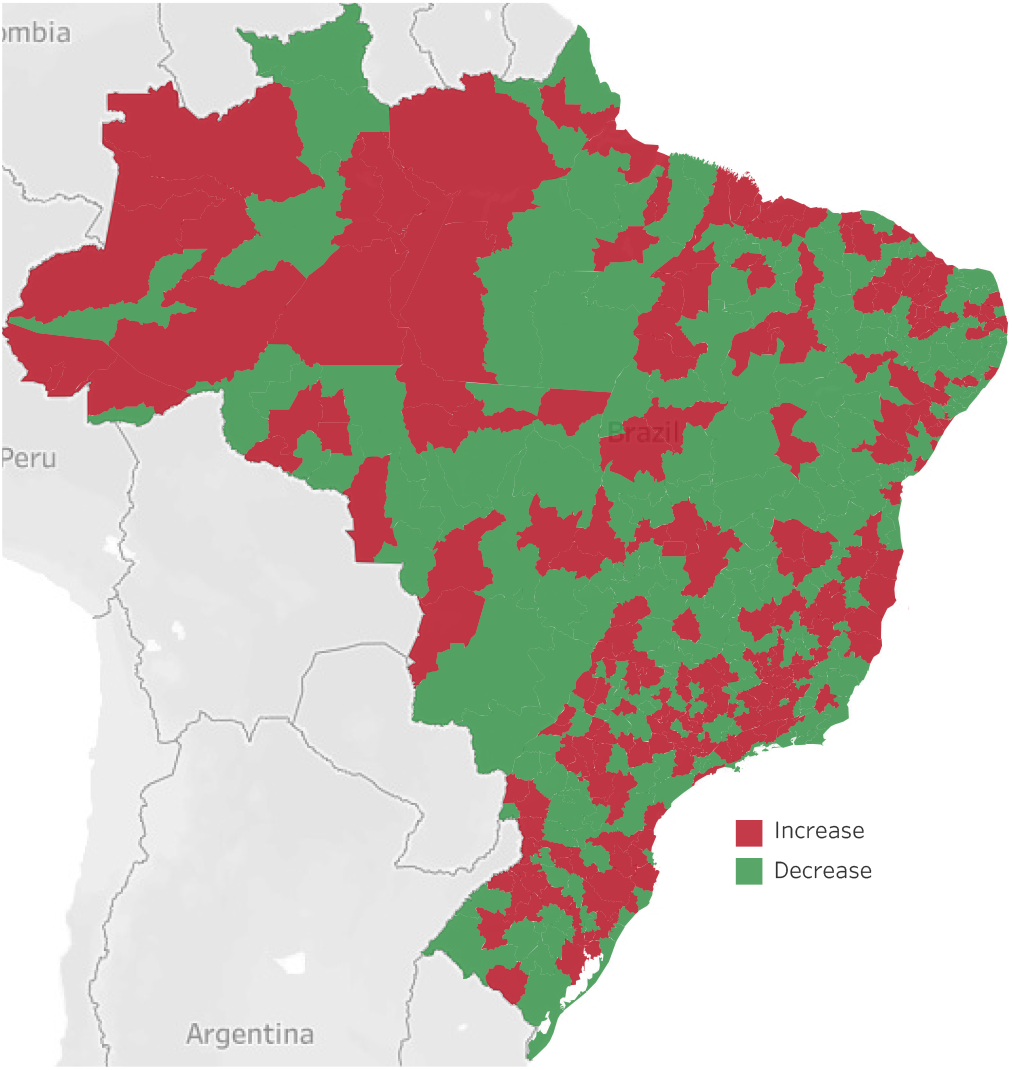
Variation in Neonatal Mortality Rate by Administrative Health Region from 2016 to 2017 – Brazil source: Public Tableau - Department of Surveillance of Noncommunicable Diseases and Diseases and Health Promotion.

In one of the few studies related to NMR forecasting in Brazil, Sergio *et. al*. [27] analyzed the annual mortality rates from infectious diarrheal diseases in children under 5 years old in Brazilian municipalities. The rates were analyzed using multilevel model, with years as first level units nested in municipalities as second level. The goal of the study was to predict yearly mortality rate by municipality. The authors conclude that there was an important difference in the pattern of mortality rate, which decreases over time, comparing the country’
s five geographic regions. However, even with this kind of study, factors associated with changes in neonatal mortality indicators in Brazil have not been fully elucidated.

On the other hand, a model that provides future neonatal mortality rates allows the anticipation of problems and demands, providing valuable information for the establishment of health policies and actions, and favoring the development of effective health promotion, care and prevention programs.

## 3. Dataset Construction

In order to build the dataset used for forecasting the neonatal mortality rate, first we construct a baseline dataset named **BRNeoDeath**^2^. From it, we constructed the **NeoDeathForecast** used in the time series model presented on this paper.

BRNeoDeath dataset has been constructed compiling data provided from two of the main sources of information related to births and deaths in Brazil: (1) SIM, which is fed using information from Declaration of Death (DO) and provide information related to death records in Brazil; and (2) SINASC, which is fed using information from Live Birth Statement(DNV) and provides demographic and epidemiological data related with the child, mother, prenatal care and childbirth [20, 3]. The association between SIM and SINASC samples was performed using NUMERODN field presented in both declarations. From the SIM, it was used the death feature to build the variable that classifies into positive (death) or negative (alive) samples, being the positive samples those comprising individuals where death occurred before the first 28 days of life.

The BRNeoDeath is similar to the dataset used by Beluzo *t*.*al* [3, 1] on their research, and has the same features, but with data from all Brazilian cities. It was constructed using live birth and death data records from Brazil between 2006 and 2016. It comprises 28,363,480 samples with the following 23 features [2]:

- **Socioeconomic and maternal conditions:** age, education, marital status and race/skin color;
- **Maternal obstetrics:** number of live births, fetal losses, number of previous gestation, number of normal and cesarean labor, estimate type and type of pregnancy;
- **Related to the newborn:** birth weight, number of pregnancy weeks, apgar index first minute, apgar index fifth minute, congenital anomaly and type of presentation of the newborn;
- **Related to previous care:** number of prenatal appointments, labor type, if the cesarean section occurred before labor began, if the labor was induced and Robson 10-groups classification

In order to prepare the data for the second major step it was necessary to preprocess **BRNeoDeath** the dataset. First each sample **BRNeoDeath** has been associated with one of the 438 administrative health regions in Brazil. This information was retrieved from one of DATASUS public repository, Territorial Units [10], and was linked using a municipal code field. Than the features originally numerical were transformed into categorical by using the approach One-hot Encoding [5].

After that, in order to build the time series dataset, each one of the new features, created by the One-hot Encoding process, were grouped by the tuple (health region, year, month) counting the number of occurrence on each features. Just for instance, consider the feature Robson Group, which is a variable that can assume 10 possibles values (categories). For this feature, the One-hot Encoding process will generate 10 new features (that will be named Robson-1, Robson-2, …, Robson-10), and after the summarizing, the feature Robson-1 represents the number of occurrences for the category 1 in Robson group feature, in a specific month and year, and for a determined health region.

After this transformation we calculate the neonatal mortality rate in this time series dataset with the count of deaths before the 28^*th*^ day of live divided by the count of live births on that specific health region and month of birth adding a new column with the variable NMR.

In addition, three more variables related to **health access** were included: number of health centers, number of neonatal incubator equipment and number of neonatal intensive care beds). These information was compiled, on monthly bases and for each health region, from another Brazilian information system, the National Register of Health Facilities (Cadastro Nacional dos Estabelecimentos de Saúde CNES), a public database containing data from all Brazilian health facilities [23].

This process allows us to transform the baseline dataset of 28,363,480 samples into a new dataset with 57,816 samples named **NeoDeathForecast**, which comprises a time series of 11 years (2006-2016), grouped by month, having 132 samples and 122 features for each one of the 438 health regions in Brazil.

## 4. Proposed Method

The mortality rate prediction can be a very powerful tool to help policy makers on their public health policies. On this section, we provide details about **NeMoR**, a new datadriven model for **Ne**onatal **Mo**rtality **R**ate forecasting. An overview of NeMoR is depicted on Figure 2, and consists of the three main steps below:

1. **Pre-processing Input Data:** the first step consists of handling the input data to deal with data gaps, which are very common in this kind of information. In problems involving time series, the occurrence of data gaps can make the forecasting unfeasible, so we propose a simple data correction to avoid this kind of problem, making possible for the NMR forecasting to be performed;
2. **Projection of Input Data Into Average NMR Space (NMRS):** after correct input data, the second step of the proposed method consists in projecting input data into a normalized feature space, which minimizes big variations along the series, and reduces noise in data. To perform this task, we take advantage of moving average approach, and propose a new method named Average NMR Space (NMRS).
3. **Forecasting NMR:** the last step of the proposed method consists in perform forecast NMR for *x* ∈ {1, 2, 3, 4} months ahead, using as input the features projected into NRMS.

**Figure 2:**
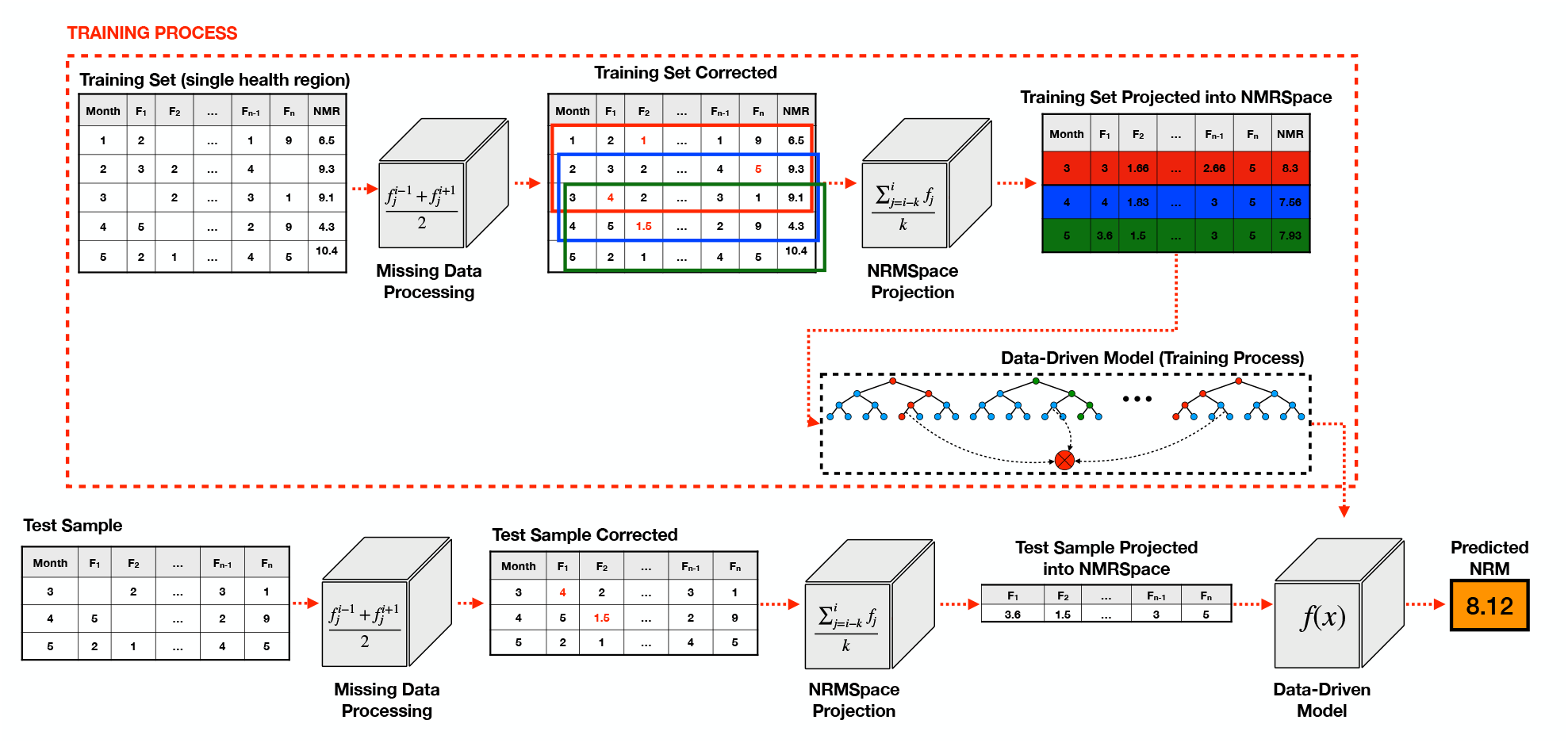
Overview of NeMoR, a new data-driven model for neonatal mortality rate fore-casting.

### 4.1. Pre-processing Input Data

The first step of the proposed method consists of filling gaps on input data for input features. This step is very important given that, on time-series forecasting it can make the method application unfeasible in a real world scenario, once missing data is a very common condition on this scenario.

Given that a sample *s*_*i*_ ∈ *S*_*R*_ is composed by the *m* features described before

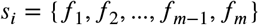

and

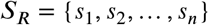

is the set of all samples in a monthly time series constructed for the health region *R*, to fill a data gap occurred in feature *f*_*j*_ in a sample *i*, proposed method uses:

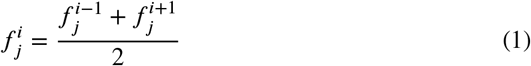

### 4.2. Features Projection Into Average NMR Space (NMRSpace)

The next step of the proposed method consists of project corrected features in a more smooth feature space, to avoid that natural noise present inn input data produce noisy and useless results. We named this new feature space as Average Neomortality Rate Space (NMRSpace).

To perform this task, we take advantage of Moving Average (MA) approach, which is largely applied on statistics to analyze data points by creating a series of averages of different subsets of the full data set. In finance, MA is also used as a stock market indicator commonly applied in technical analysis [8]. By calculating the MA, the impacts of random and short-term fluctuations on the NMR over a specified timeframe are mitigated, producing better quality features. The same idea can be applied over the other features in proposed method, in a way to smooth noise and produce better quality features.

To project an input feature into NMRSpace,the proposed method takes the arithmetic mean for each feature. Equation 2 depicts how to calculate a projected sample:

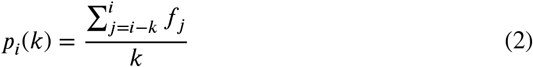

This way, to calculate the *i*^*th*^ projected sample *p* is necessary to use features from time stamps *i, i* − 1, …, *i* − *k* associated with features measurements {*f*_*i*_, *f*_*i−1*_, …, *f*_*i−l*_} where *k* < *i*.

### 4.3. Forecasting NMR

The last step of the proposed method consists of predicting NMR for *d* months ahead, where *d* ∈ {1, 2, 3, 4}, using a regression model and features projected on NMRSpace.

NeMoR uses Extreme Randomized Trees, also named Extra-Trees, as its regression method. The Extra-Trees algorithm has been proposed by Geurts et al. [11] in 2006. The algorithm is very similar to random forest algorithm, but it applies a simpler algorithm to construct the decision trees used as members of the ensemble. Another advantage of the algorithm is that it is a limited number of key hyperparameters and sensible heuristics for configuring hyperparameters.

As described by Geurts et al. [11], the algorithm builds an ensemble of unpruned decision or regression trees according to the classical top-down procedure. Its two main differences with other tree-based ensemble methods are that it splits nodes by choosing cut-points fully at random and that it uses the whole learning sample (rather than a bootstrap replica) to grow the trees.

The main hyperparameters on Extra-Trees are:

- *K*: represents the number of attributes randomly selected at each node. Along training process, *K* determines the strength of the attribute selection process;
- *n*_**min**_: represents the minimum sample size for splitting a node. It is also responsible by algorithm strength relative to averaging output noise;
- *M*: represents the number of trees of this ensemble. The main function of *M* is determining the strength of the variance reduction in the ensemble model aggregation.

## 5. Experiments and Results

In order to evaluate the proposed method on different scenarios, this section presents different rounds of experiments along with details for each experiment.

### 5.1. Computational Environment Setup

This method was implemented in Python Language (version 3.7) and the most known libraries in Python used in Data Science, such as Scikit-Learn (0.23.1), Pandas (version 1.0.4) and MatplotLib (version 3.2.1). All the experiments have been performed in a computer running Ubuntu 19.04 (64 bits).

### 5.2. Dataset, Metrics, and Validation Protocol

As described on Section 3, we constructed a time series dataset focused on NMR prediction. However, given the high number of health regions (438), we evaluated the proposed method in the São Paulo health region, given the better data quality for this region^3^ [2, 3].

In order to evaluate the quality of the proposed method, we use different error metrics as Mean Squared Error (MSE), Mean Absolute Error (MAE), Root Mean Squared Error (RMSE), Mean absolute percentage error (MAPE) [22]. Furthermore, we depict time series curves comparison (predicted *υs*. ground truth) for each evaluated scenario.

Given that the proposed dataset contains a time series of 132 months, as a validation protocol we combine a sliding window and leave-one-out validation protocols. This way, we use a training set window of 60 months (e.g. from month 1 to 60) to train the model, and *x* months ahead to test it (in case of predictions 2 months ahead for example, we use month 62). Then, we move our window one position to the right, keeping 60 months on training set (e.g. from month 2 to 61), testing the model on month 63, etc. This way, reported metrics represent the average of all 60 training/test rounds.

### 5.3. Features Selection For NMR Forecasting

From the entire dataset constructed, as described on Section 3, our method performs NMR forecasting based on five features:

- **Mortality:** Neonatal Mortality Rate.
- **UBS:** Number of Health Centers.
- **Incubator:** Number of Neonatal Incubator Equipment.
- **Education:** Education of the mother in years completed (8 to 11 education years).
- **NeoUnit:** Number of Neonatal Intensive Care Unit Beds.

The idea behind using these features is finding features where it is possible for policy makers to take necessary actions to decrease a probable rise on NMR, once they have mechanisms to intensify campaigns to reduce this rates.

As features of the selecting method, a correlation analysis of each variable with NMR was performed, none of the features reached a relevant correlation value (all below 0.3), therefore the features were selected based mainly on the problem domain. Figure 3 depicts results for features achieving bigger correlation values. Thus, the five features selected to be taken as input for NeMoR method were chosen because they are associated with neonatal mortality, and which are also subject to interference by public policies, i.e., in some way, could be improved by public health investments (e.g. the labor type, prenatal, number of centers, etc.),

**Figure 3:**
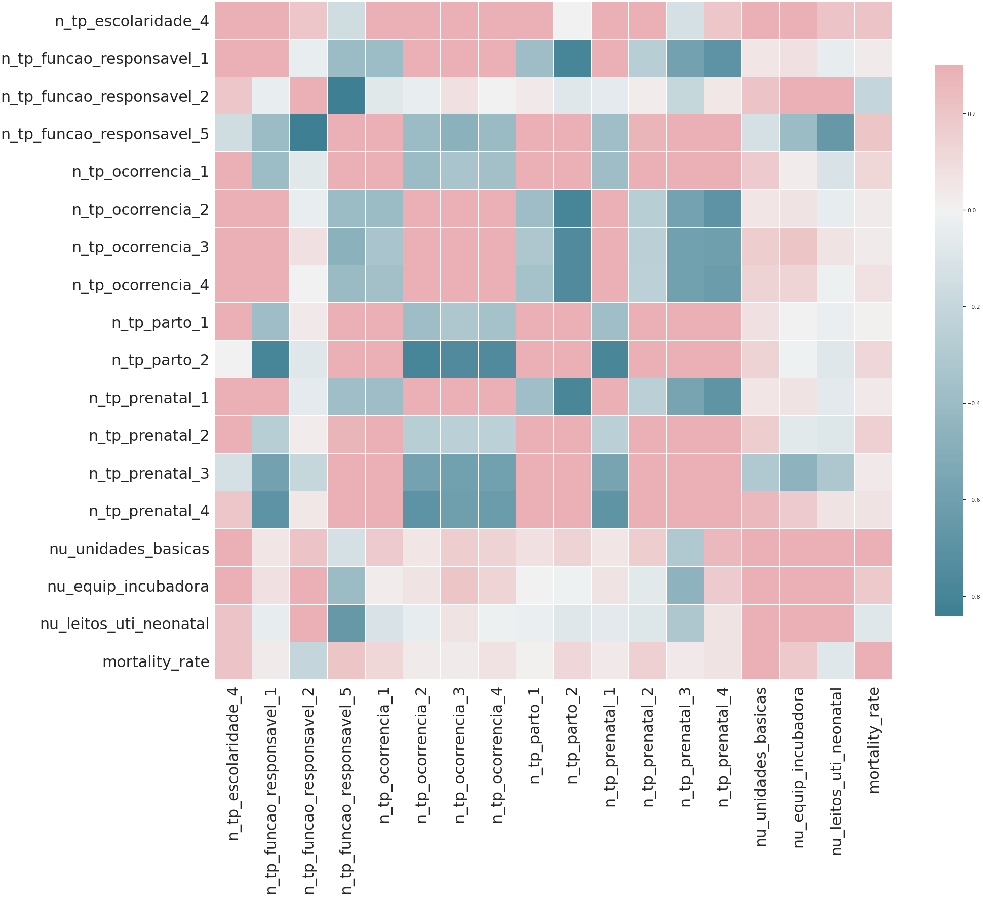
Correlation matrix for the most important features.

In our dataset, each one of these features describes the occurrences number of that specific feature in an specific month for one specific health region. So, one sample for health region São Paulo in May 2009 will represent, for that specific month, the mortality (NMR) registered (Mortality), the number of health centers per region (UBS), the number of neonatal incubator equipments located in the region (Incubator), the number of neonatal intensive care unit beds reported in the region (NeoUnit) and the number of women with education level between 8 and 11 years, who gave birth (Education) in that month.

### 5.4. Experiment #1: Evaluating Different Regression Models

As previously mentioned, our method has been validated using São Paulo time series, mainly because of data quality but also because it is the biggest metropolis in Brazil. On this first round of experiments, we evaluated different regression models for NeMoR using different forecasting windows (1, 2, 3, and 4 months ahead). Besides Extra-Trees, we also evaluated 3 additional regression models: XGBoost Regressor [6] (XGB), Gradient Boosting Regressor (GBR), and Lasso [13]. Figures 4a, 4b, 4c, and 4d depicts, respectively, results for forecasting window of 1, 2, 3 and 4 months ahead. Additionally, Table 1 depicts evaluation metrics for each regression model.

**Figure 4:**
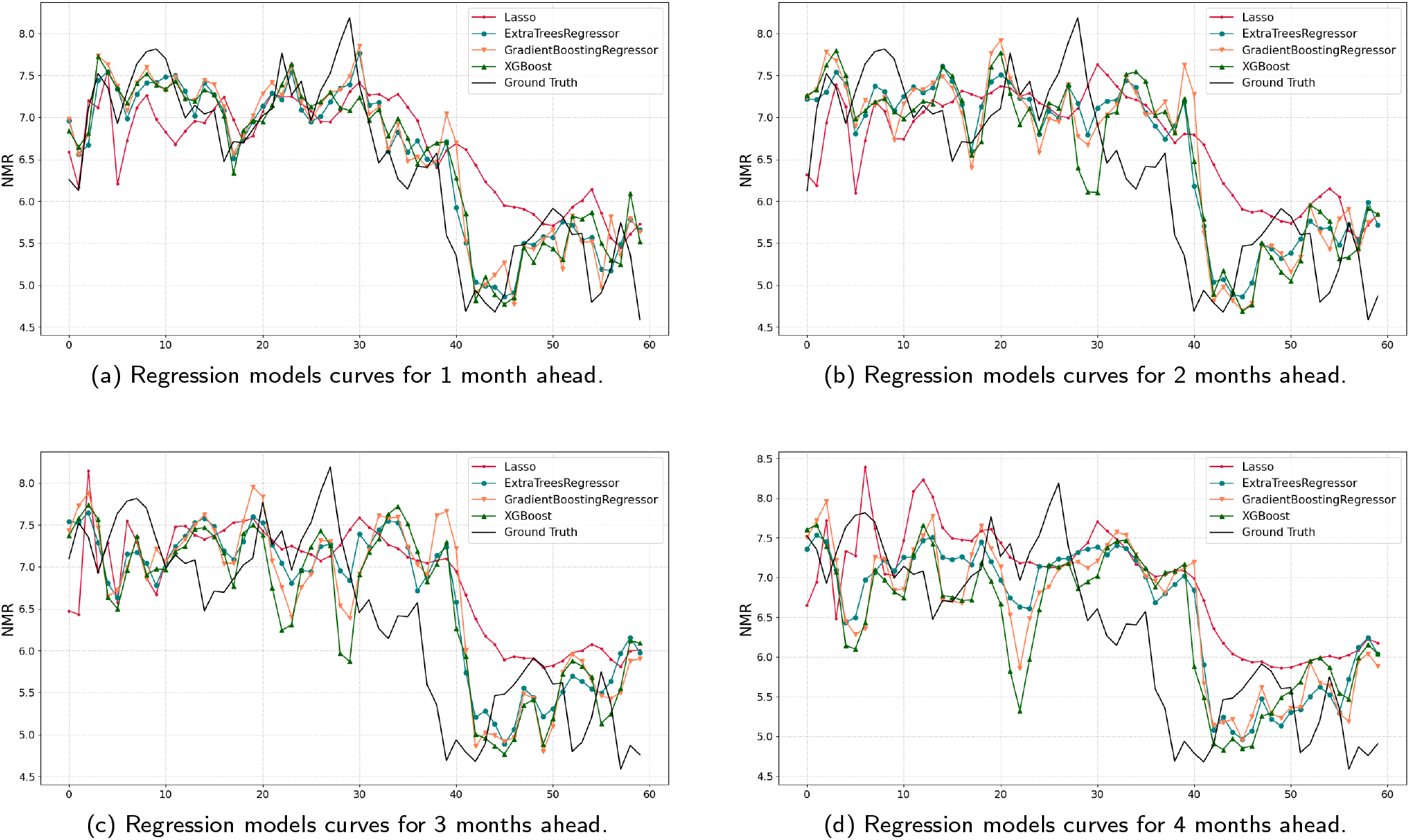
Prediction curves for evaluated regression models using different prediction windows.

**Table 1:**
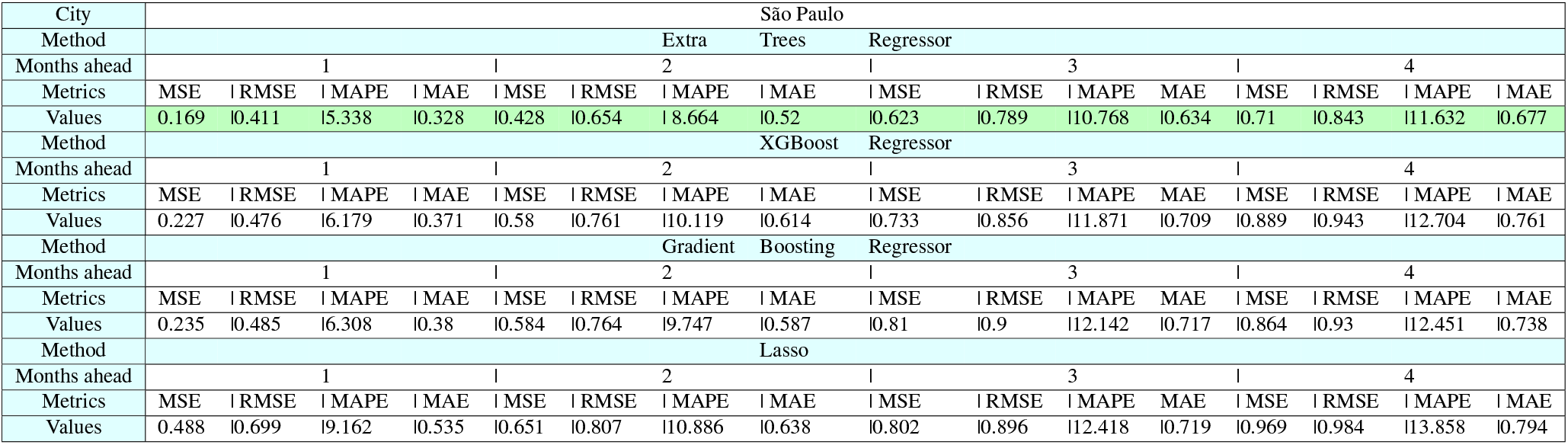
Metrics for for 1, 2, 3 and 4 ahead for each regression model evaluated.

As it can be observed from Figures, the linear model (Lasso) presents worse results for all 4 scenarios evaluated. By a visual inspection, GBR, XGB and Extra-Trees present very similar results, depicting a good accuracy on time series behavior (curve shape) and prediction (distance between curves), mainly on 1 and 2 months ahead. On the other hand, for 3 and 4 months, these methods hold accuracy on series behavior but lost accuracy on prediction (curves more distant from ground truth curve). This type of behavior can be of great use to policy makers in the planning of public health policies since it allows them to analyze trends of rise and fall in NMR.

When evaluating numerical metrics, Extra-Trees and XG-Boost performs very closer to each other, but Extra-Trees still presents superior results.

Even without a perfect accuracy, NeMoR can be very useful to predict the NMR movement and tendency, being a very useful tool for policy makers.

### 5.5. Experiment #2 - Fine Tuning ExtraTree Model

From a previous experiment, we selected the ExtraTree regression model to take place as the last step of the proposed method. Then, on this section we performed an experiment to evaluate the gain obtained by a fine-tuned ExtraTree model. The parameters evaluated and it’s respective domain are:

- nestimators: [50, 150, 250, …, 750]
- minsamplesleaf: [1, 2, …, 6]
- minsamplessplit: [2, 3, …, 5]
- criterion:mse, friedmanms, mae
- maxdepth: [1, 2, …, 5, unlimited]
- minweightfractionleaf: [0.1, 0.2, …, 0.5]
- randomstate: [1, 2, …, 5, unlimited]
- minimpuritydecrease: [0.0, 0.5, …, 2.0]
- minimpuritysplit: [0.0, 0.5, …, 2.0, unlimited]
- maxleafnodes: [2, 3, …, 5, unlimited]

Figures 5(a)-(d) depicts results comparison between curves before and after ExtraTree fine-tuning. From these results, we concluded that only the parameter **nestimators** improves results when compared to default values. The best number of estimators obtained for each evaluated scenario are:

- 1 Month Ahead - estimators = 70
- 2 Months Ahead - estimators = 120
- 3 Months Ahead - estimators = 80
- 4 Months Ahead - estimators = 120

**Figure 5:**
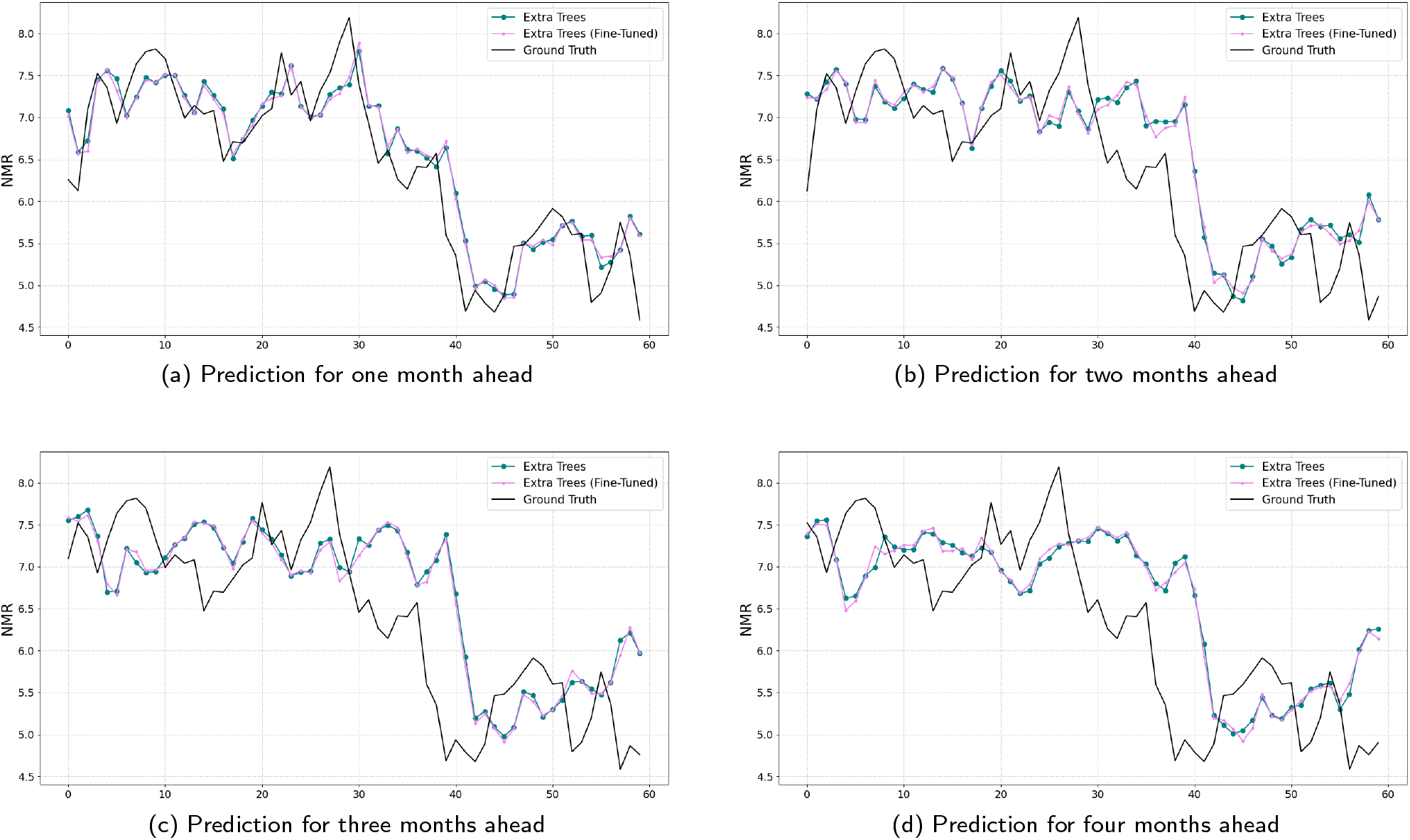
Curves for the proposed method, using fine-tuned ExtraTree model as a regression model.

Its worth to highlight here that, even on the best fine-tuned model, the statistical difference between the model before and after fine-tuning is not significant, as presented on Table 2 that depicts metrics comparison for the same scenario (before and after fine tuning).

**Table 2:**
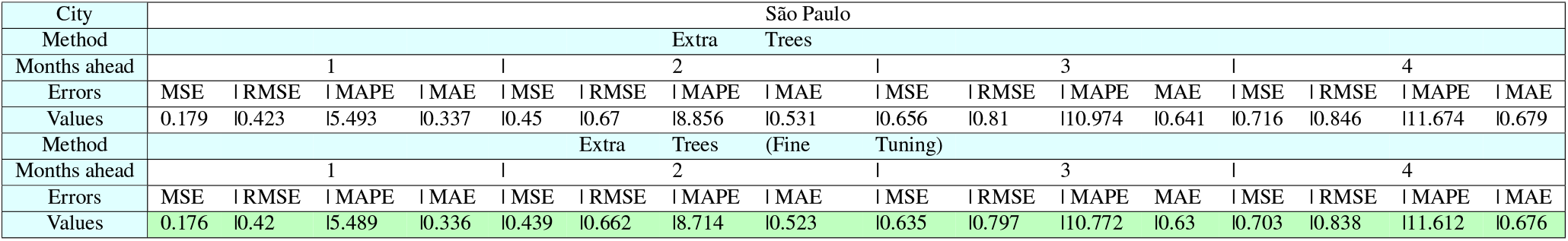
Predictions for the next four months, before and after fine-tuning.

The rest of the experiments on this paper have been performed using fine-tuned ExtraTree model.

### 5.6. Experiment #3: The Importance of NMRSpace

The third round of the experiment is focused on exposing the importance of NMRSpace for the NeMoR accuracy.

To propose NMRSpace, we started using a visual inspection of NMR time series for São Paulo city, as depicted on Figure 6. Then, we calculated the univariate distribution for the NMR time series, as depicted on Figure 7.

**Figure 6:**
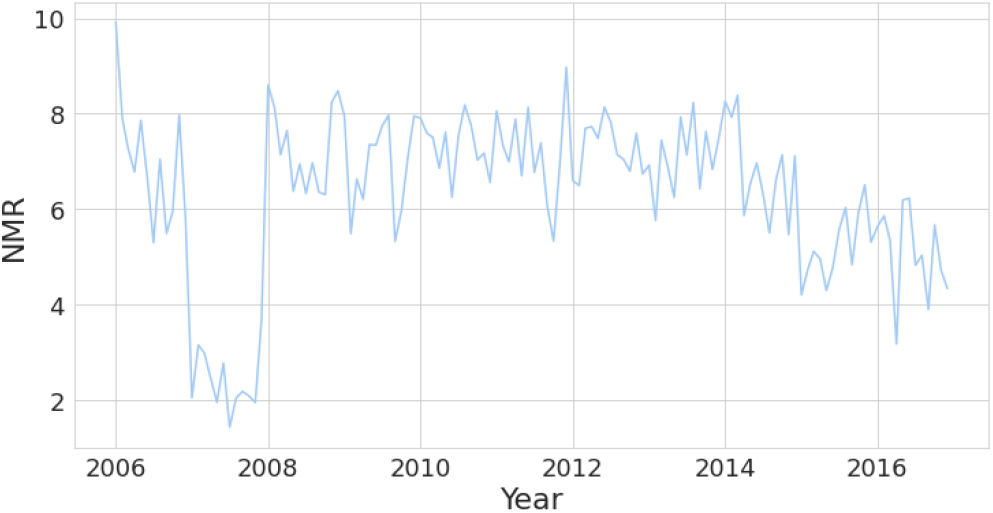
Overview of NMR from 2006 to 2016 for São Paulo city.

**Figure 7:**
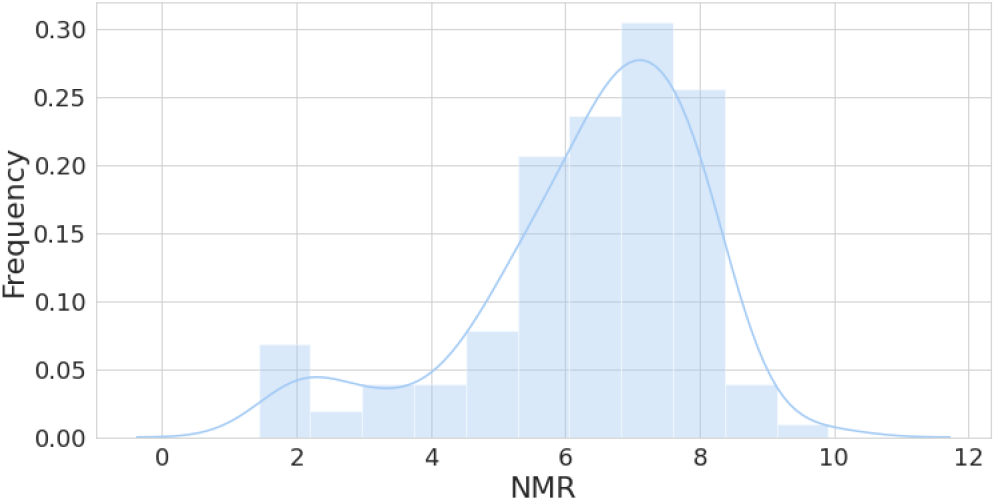
Univariate distribution of NMR time series for São Paulo city.

As it can be observed, the data presents a Gaussian distribution, which indicates a non-stationary pattern, making it hard to apply classical prediction methods over the data without any transformation. By using the augmented Dickey–Fuller test (ADF) [7], which tests the null hypothesis that a unit root is present in a time series sample, we achieved a value of −3.54. Values lower than 0 points toward less stationary time series. Furthermore, the time series achieved a p-value[30] of 0.007, which reinforce our hypothesis of non stationary series (values lower 0.05 are considered non stationary).

All these evidences point toward the necessity of a data transformation to better fit, not just NMR, but also the time series of other selected features. Then, we evaluated two scenarios: when using and when not using NMRSpace projection for NeMoR classification. Table 3 depicts the metrics for each scenario evaluated. Its not difficult to realize that prediction using NMRSpace produce better metrics than metrics produced by NeMoR when applied over raw data (without data projection).

**Table 3:**
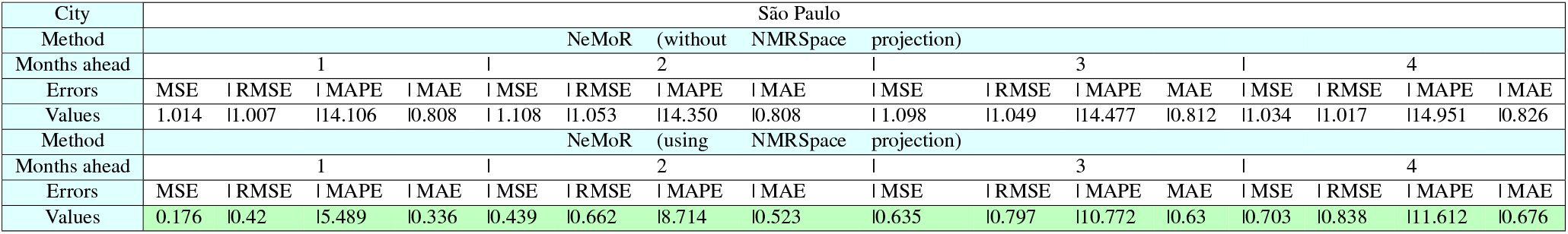
Metrics comparing NeMoR results using, or not, NMRSpace features projection.

Furthermore, we did evaluate different values for *k* parameter in NMRSpace projection, defined on Equation 2. Table 4 presents NeMoR metrics when varying *k* in NMRSpace.

**Table 4:**
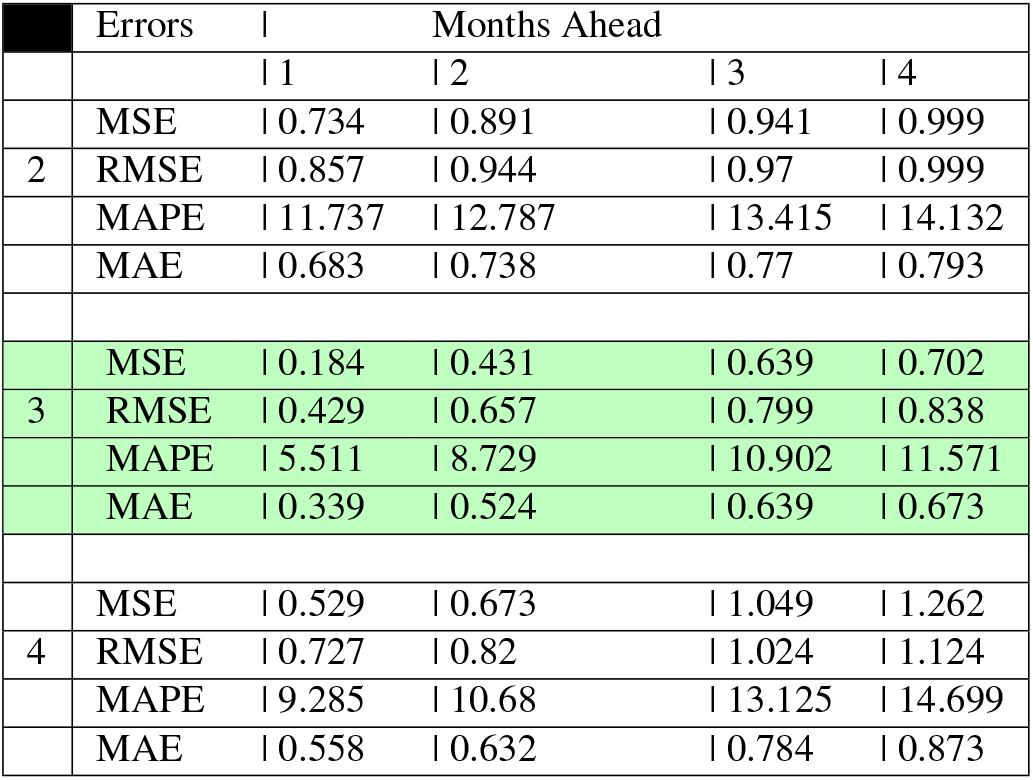
Results for NeMoR when varying the *k* parameter on NMRSpace.

As it can be seen, a value of *k* = 3 provides better results when compared to other values with a significant difference in results of all evaluated scenarios (1, 2, 3, and 4 months ahead).

## 6. Discussion

Neonatal mortality indicators are very important to public health systems evaluation and are strongly related to the quality of prenatal and childbirth care. To achieve reduction of neonatal mortality it is necessary to have a deep understanding of the levels and trends in neonatal mortality. Important achievements have been achieved around the world, but improvements are still needed in some places, like in Brazil. In this context our paper provides knowledge that may help a deep understanding of the factors related with neonatal mortality and contribute in the prevention by providing a new forecasting method based on data extraction knowledge and machine learning techniques.

On this paper we developed a data-driven model to predict the neonatal mortality for São Paulo city in the next one, two, three and four months ahead. Our dataset contains information from all Brazilian administrative health regions, over 11 years (2006 - 2016). The new time series dataset built on this paper, NeoDeathForecast, is an important contribution as resources for researches specially on public health policies and demography subjects for NMR prediction problems lems.

By combining the features: (a) previous NMR, (b) number of health care units, (c) number of neonatal incubators in region, (d) number of mothers having years of schooling between 8 to 11, and (e) number of newborn intensive care units in the region, our method NeMoR was able to achieve a forecasting of NMR with an a MSE lower than 0.18, on the best experiment setup. Although with a high value for MSE, the NMR forecast results of NeMoR method still may be useful providing a way for policy makers to anticipate trends on NMR curves, being an important tool for planning public health policies. The feature selection was guided by factors that could mitigate neonatal mortality and which policy makers could directly act and based on correlation value with NMR.

In contrast to classical statistical models, NeMoR is a data-driven method, and relays on knowledge learned from a high-volume dataset, an important factor for the method validation. Besides that, four different ML regression models were experimented, Extra Tree presenting best results between then, as well as a hyperparameters search step as model fine tune.

## Data Availability

Dataset and source codes used on the experiments will be made public under paper acceptance.

## Footnotes

1 The health regions is a continuous geographic space constituted by a grouping of neigh-boring municipalities in order to integrate the organization, planning and execution of actions and health services.

2 Additional details about BR Neo Death are described in a paper under evaluation.

3 All the results reported on this paper can be easily reproduced for other regions, once dataset and source will be made public under paper acceptance

